# The Cost of Distrust: Governance Perceptions, Risk, and COVID-19 Vaccine Uptake in Africa—Evidence from the Afrobarometer Survey

**DOI:** 10.1101/2025.11.26.25341108

**Authors:** Richmond Balinia Adda, Conrad Pwayirane

## Abstract

**Background:** The stark heterogeneity in COVID-19 vaccine uptake across Africa cannot be fully explained by individual-level factors or access alone. This study investigates the crucial, yet underexplored, role of political and governance perceptions in shaping health behavior. We examine how citizen perceptions of governance, specifically institutional trust, perceived corruption, and satisfaction with health services, influence COVID-19 vaccination uptake in Africa. We further test whether perceived government preparedness, as a measure of institutional risk mitigation, mediates this relationship.

**Methods:** We analyzed cross-sectional data from 53,444 respondents across 39 African countries (Afrobarometer Round 9, 2021-2023). Using multilevel logistic regression and generalized structural equation modelling, we assessed associations while controlling for sociodemographic and accounting for country-level clustering.

**Results:** Our analysis reveals three key patterns. First, institutional trust has a non-linear relationship with vaccination. Respondents with **“just a little”** trust had **∼28.6%** uptake, compared to **23.1%** for those with **“a lot”** of trust. Second, corruption perceptions exhibited a dose-response effect; a one-unit increase on the corruption scale was associated with a **1.8%**-point reduction in vaccination probability (**p<0.001**). Third, we identified a complacency effect: a one-unit increase in perceived government preparedness was linked to **20.2%** lower odds of vaccination (**p<0.001**). Substantial heterogeneity existed: the effect of institutional trust was twice as strong in urban areas (**OR=1.050**) than in rural areas (OR=1.025). Multilevel modelling showed that country-level factors accounted for 29% of the total variance in vaccination behavior.

**Conclusion:** Governance perceptions are fundamental determinants of health behavior in Africa. The robust negative effect of corruption and the counterintuitive complacency effect underscore that building trustworthy, transparent governance is a public health imperative. However, communicating institutional capability requires careful framing to avoid undermining personal responsibility.

## Introduction

The stark heterogeneity in COVID-19 vaccine uptake across Africa presents a critical puzzle for public health-related Sustainable Development Goals in Africa (World Health Organization, 2021). Despite widespread availability, acceptance varied dramatically between and within countries, suggesting factors beyond mere access or individual knowledge were at play (GBD 2019 Diseases and Injuries Collaborators, 2020). This study argues that resolving this puzzle requires understanding citizens’ perceptions of their governments. We examine how institutional trust, perceived corruption, and satisfaction with health services shaped vaccination behavior during the pandemic. The COVID-19 pandemic has further highlighted the pivotal role of population-level behavior change in determining public health outcomes, making the understanding of health behavior determinants an urgent priority for both research and policy.

Despite extensive awareness campaigns and health education initiatives, adherence to recommended health behaviors often remains suboptimal across African contexts (Dzinamarira et al., 2021). This persistent gap between knowledge and action suggests that factors beyond information and awareness are at play. Traditional health behavior theories, particularly the Health Belief Model (Rosenstock, 1974) and Theory of Planned Behavior (Ajzen, 1991), have emphasized individual-level psychological determinants such as perceived susceptibility, severity, benefits, and barriers. While these frameworks provide valuable insights, they often overlook the crucial role of broader political and institutional contexts in shaping individual health decisions (Blair, Morse, & Tsai, 2017).

The governance-health nexus represents a critical but understudied dimension of health behavior adoption. Distrust in government institutions can lead to skepticism toward official health information and recommendations (Devine et al., 2020). Perceived corruption creates a sense of unfairness and reduces willingness to comply with state-led initiatives (Rothstein & Uslaner, 2005). Dissatisfaction with health services undermines confidence in the entire health system’s efficacy, creating barriers to preventive care utilization (Kruk et al., 2018). These governance perceptions may operate through multiple pathways: directly by influencing the perceived legitimacy of health directives, and indirectly by shaping risk perceptions and behavioral intentions (Bollyky et al., 2022).

While the relationship between political trust and health behavior has been examined in high-income countries (O’Keefe, 2021), it remains under-explored in the diverse African context, where governance challenges are often more acute and health systems face greater resource constraints. The Afrobarometer dataset provides a unique opportunity to fill this gap, offering comparable data across multiple African countries on both governance perceptions and health behaviors (Mattes, 2020).

This study addresses three research questions:

1. To what extent are trust in government/health systems, perceived corruption, and satisfaction with health services associated with adherence to preventive health behaviors?
2. How does perceived health risk mediate the relationship between governance perceptions and health behavior?
3. Do these relationships vary by country or individual socio-demographic characteristics?

By examining these questions, this research contributes to both theoretical understanding of health behavior determinants and practical strategies for improving public health outcomes in Africa.

### Literature Review & Theoretical Framework

#### Health Behavior Theories: Foundations and Limitations

The Health Belief Model (HBM) and Theory of Planned Behavior (TPB) represent two dominant frameworks for understanding health behavior adoption. The HBM posits that individuals are more likely to engage in preventive health behaviors when they perceive themselves as susceptible to a condition, believe the condition has serious consequences, recognize the benefits of action, and identify few barriers to action (Rosenstock, 1974; Champion & Skinner, 2008). The TPB extends this framework by incorporating subjective norms and perceived behavioral control as additional determinants of behavioral intentions and actions (Ajzen, 1991; McEachan et al., 2011).

While these theories have demonstrated utility across diverse health contexts, they have been criticized for their predominantly individualistic focus and limited attention to structural and political determinants (Blair et al., 2017). Recent research suggests that institutional and political factors may significantly moderate the relationships proposed by these traditional models (Bollyky et al., 2022).

#### Trust and Health Compliance: The Institutional Dimension

Political and institutional trust has emerged as a critical factor in public compliance with health recommendations. Drawing on theories of political legitimacy (Levi & Stoker, 2000), researchers have found that trust in government institutions predicts compliance with public health directives during emergencies, including vaccination uptake and adherence to preventive measures (Devine et al., 2020; O’Keefe, 2021). The mechanism appears to operate through multiple pathways: trusted institutions are perceived as more credible information sources, their directives are viewed as more legitimate, and their recommendations inspire greater confidence in their efficacy (Blair et al., 2017).

In African contexts, where colonial legacies and post-independence governance challenges have often created trust deficits, the relationship between institutional trust and health behavior may be particularly salient (Logie et al., 2021). However, this relationship remains inadequately studied across the continent’s diverse political landscapes.

#### Corruption and Public Health: Erosion of Trust and Legitimacy

Corruption perceptions represent a significant barrier to public health efforts worldwide. Research indicates that perceived corruption erodes institutional trust, reduces the legitimacy of government directives, and creates cynicism about the fairness and effectiveness of public programs (Rothstein & Uslaner, 2005; Holmberg & Rothstein, 2011). In health contexts, corruption perceptions have been linked to reduced utilization of health services, lower vaccination rates, and decreased compliance with public health guidelines (Kruk et al., 2018; Gaitán-Rossi et al., 2021).

The mechanisms linking corruption to health behavior include both practical concerns (e.g., diversion of resources reducing service quality) and psychological effects (e.g., reduced motivation to comply with directives from corrupt institutions) (Rothstein, 2011). In many African countries, where corruption is frequently identified as a major public concern, these effects may be particularly pronounced (Mattes, 2020).

#### Service Delivery Satisfaction: From Experience to Expectation

Satisfaction with health services represents a crucial link between past experiences and future health behaviors. Drawing on models of service quality and patient satisfaction (Andaleeb, 2001), research has demonstrated that negative experiences with health services reduce future care-seeking and adherence to medical advice (Kruk et al., 2018). This relationship operates through both cognitive pathways (e.g., expectations of poor-quality care) and affective pathways (e.g., frustration or disillusionment with the health system).

In contexts of limited resources and infrastructure, such as many African health systems, service quality issues may be particularly salient in shaping health behaviors (Amporfu et al., 2022). Satisfaction with government performance in health service delivery may serve as a proxy for broader confidence in the health system’s capacity and effectiveness.

#### Theoretical Synthesis and Conceptual Framework

Integrating these diverse literatures, we propose a conceptual framework that positions governance perceptions as fundamental determinants of health behavior, operating both directly and through mediating psychological mechanisms (**Figure 1**). This framework builds on established health behavior theories while incorporating crucial political and institutional dimensions often overlooked in traditional models.

**Figure 1.**
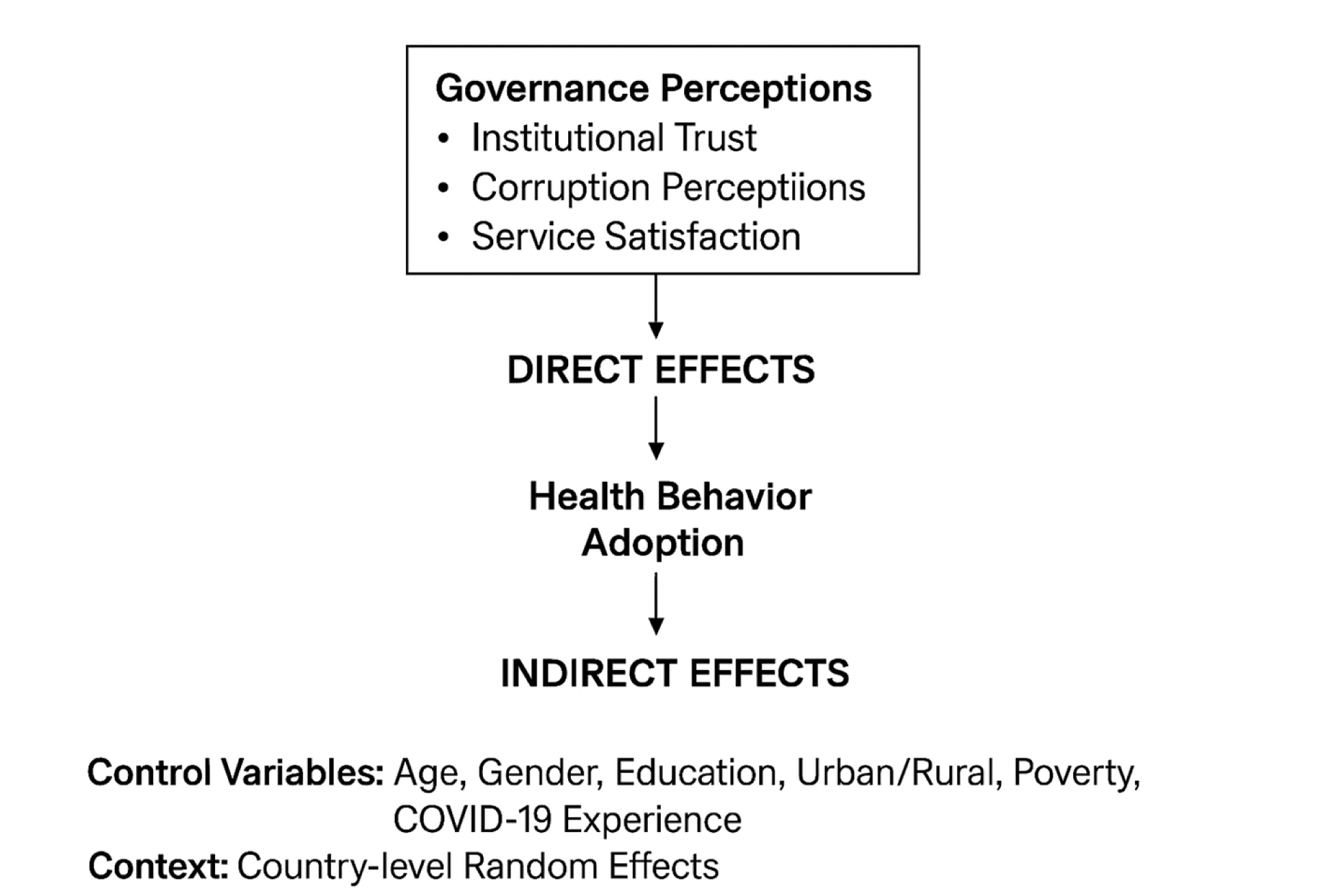
Conceptual Framework of Governance Perceptions and Health Behavior.

This framework posits that governance perceptions influence health behavior through two primary pathways. First, direct effects operate through the perceived legitimacy of health directives and confidence in institutional capabilities. Second, indirect effects operate through risk perception mechanisms, where governance perceptions shape assessments of threat severity and susceptibility. The framework also acknowledges the importance of controlling for individual-level characteristics and accounting for country-level contextual factors.

By testing this framework across diverse African contexts, this study addresses critical gaps in understanding how political and institutional factors shape health behavior decisions, with important implications for both theory and public health practice.

### Methodology

#### Data Source and Study Population

This study utilised data from Afrobarometer Round 9, collected between 2021 and 2023 across 39 African countries with a total sample of 53,444 respondents. This data, used for this analysis, was accessed and downloaded for research purposes in November 2025. The dataset is a public-use file from which all direct identifiers have been removed; therefore, the authors had no access to information that could identify individual participants at any stage of this research. The Afrobarometer represents a pan-African, non-partisan research network that conducts high-quality, nationally representative surveys on public attitudes toward governance, economic conditions, and social issues. The survey employs a rigorous multi-stage stratified random sampling design that ensures representativeness at both national and sub-national levels, while simultaneous data collection across multiple countries enables robust comparative analysis.

The analytical sample for this study ranged from 49,042 to 49,889 respondents across different model specifications, reflecting listwise deletion of missing data. Vaccination intention models utilized smaller samples (n=21,598) due to higher non-response rates for hypothetical behavioral questions. All analyses incorporated sampling weights to account for complex survey design and ensure population representativeness.

#### Study Design

We employed a cross-sectional observational design with multilevel modelling to account for the hierarchical structure of individuals nested within countries. While cross-sectional data preclude causal inference, the large sample size and comprehensive covariate adjustment enabled robust examination of associations between governance perceptions and health behavior. While this design precludes definitive causal inference, the large, representative sample and comprehensive control for confounders allow for robust examination of the associations between governance perceptions and health behavior.

#### Variables and Measurement

##### Dependent Variables

Our primary outcome was COVID-19 vaccination behavior, measured using the question: “Have you received a COVID-19 vaccination?” (1=Yes, 0=No). We selected this behavioral measure due to its stronger ecological validity and reduced susceptibility to social desirability bias compared to intention measures. Secondary analysis examined vaccination intention using the question: “How likely are you to get a COVID-19 vaccine?” measured on a 4-point scale from “Very unlikely” to “Very likely”, allowing investigation of the well-documented intention-behavior gap.

##### Independent Variables

Governance perceptions were operationalized through three key constructs. Institutional trust was measured using a composite index (Cronbach’s α=0.84) comprising five items assessing trust in the president, parliament, local government council, police, and courts. Responses were recorded on a 4-point scale from “Not at all” to “A lot” and averaged, with higher scores indicating greater trust. This multidimensional approach captures the diffuse support for political institutions that underpins policy compliance.

Corruption perceptions were measured using the question: “Over the past year, has the level of corruption in this country increased, decreased, or stayed the same?” recoded to a 4-point scale. This measure captures perceptions of systemic corruption, which prior research links to reduced compliance with public health directives.

Health service satisfaction was assessed using the question: “How well or badly would you say the current government is handling improving basic health services?” on a 4-point scale from “Very badly” to “Very well”. This construct reflects evaluations of government performance in healthcare delivery, a key dimension of political accountability.

##### Mediating Variable

Perceived Government Preparedness was operationalized as a proxy for institutional risk mitigation capacity, using the question: “How well do you think the government could prepare for future health emergencies?” recoded to a 4-point scale. We theorize that in contexts of limited health literacy, perceptions of government capability may serve as a powerful heuristic, influencing an individual’s perceived need for personal protective action, potentially inducing complacency.

##### Control Variables

We included comprehensive demographic and socioeconomic controls established in health behavior literature: age (categorized as 18-35, 36-55, 56+), gender, education level, urban-rural residence, and the Lived Poverty Index (a validated composite measure of material deprivation). Final models also controlled for personal COVID-19 experience, as direct health experiences strongly influence preventive behavior.

##### Analytical Strategy

Our analytical approach followed a sequential multi-method framework. Preliminary analyses included descriptive statistics to characterize the sample and bivariate analyses (t-tests, chi-square tests, ANOVA, and correlations) to examine initial relationships and identify potential multicollinearity issues.

Multivariate analysis employed four sequential logistic regression models to examine the governance-behavior relationship while progressively adding control variables. The sequential approach followed established practice for building multivariate models, with Model 1 using categorical trust measures, Model 2 employing the continuous trust index, Model 3 examining vaccination intentions using ordered logistic regression, and Model 4 incorporating personal COVID-19 experience.

Recognising the hierarchical structure of the data, we specified a mixed-effects logistic regression model with random intercepts for countries. This approach accounts for unobserved country-level heterogeneity and provides more accurate standard estimates. The significant random effects variance component confirmed the necessity of this approach.

We employed generalized structural equation modelling to test whether risk perception mediates the relationship between institutional trust and vaccination behavior. This method accommodates binary outcomes and mediators while providing direct and indirect effect estimates, making it particularly suitable for testing complex mediation pathways with categorical variables.

Subgroup analyses were conducted through stratification by gender and urban-rural residence to examine effect heterogeneity, aligning with growing recognition that health behavior determinants may operate differently across population subgroups.

Comprehensive model diagnostics included marginal effects analysis to interpret substantive significance, variance inflation factors to check multicollinearity, and likelihood ratio tests to compare model fit.

##### Ethics Statement

This study utilised secondary, anonymised data from the Afrobarometer Round 9 survey. The Afrobarometer network secured ethical approval for the survey from the appropriate institutional review boards in all participating countries. All respondents provided written informed consent prior to participation, following established ethical protocols. As this study involved analysis of existing, de-identified public-use data, it was exempt from further ethical review.

##### Patient and Public Involvement

This research was conducted using secondary, de-identified data from the Afrobarometer public opinion survey. Patients or the public were not involved in the design, conduct, reporting, or dissemination plans of our specific analysis.

##### Analytical Software

All analyses were conducted using Stata Version 17.0, employing specialized packages for complex survey analysis, multilevel modelling, and structural equation modelling. Statistical significance considered at 95% confidence level.

## Results

### Sample Characteristics and Descriptive Statistics

The analytical sample comprised 53,348 respondents from the Afrobarometer Round 9 survey. As shown in **Table 1**, 57.1% of respondents reported having received COVID-19 vaccination, while 42.9% remained unvaccinated. Among those expressing vaccination intentions (n=22,598), the distribution was relatively balanced across likelihood categories, with a mean intention score of 2.76 (SD=1.22) on a 4-point scale.

**Table 1:** Descriptive Statistics of Key Study Variables (N=53,348)

| Variable | Mean | SD | Min | Max | n |
| --- | --- | --- | --- | --- | --- |
| Vaccinated | 0.57 | 0.49 | 0 | 1 | 53,348 |
| Vaccine Intention | 2.76 | 1.22 | 1 | 4 | 22,598 |
| Trust Index | 4.18 | 7.39 | 0 | 94 | 53,355 |
| Corruption Perception | 1.77 | 1.33 | 0 | 3 | 51,586 |
| Health Service Satisfaction | 1.20 | 0.96 | 0 | 3 | 52,832 |

The trust index, computed across five institutional domains (president, parliament, local government, police, and courts), demonstrated substantial variation (Mean=4.18, SD=7.39), indicating heterogeneous levels of institutional trust across the sample. Perceptions of corruption were moderately high (Mean=1.77, SD=1.33 on a 0-3 scale), while satisfaction with health services averaged 1.20 (SD=0.96) on a 0-3 scale.

### Bivariate Analyses

#### Bivariate Associations Between Institutional Trust and Vaccination Behavior

We observed significant associations between institutional trust and vaccination status across multiple domains (**Table 2**). Trust in the president showed a non-linear relationship with vaccination (χ²=149.46, p<0.001). Contrary to expectations, the highest vaccination rates were observed among those reporting “just a little” trust (28.6%), followed by “somewhat” trust (21.5%), “a lot” of trust (23.1%), and “not at all” (26.9%). This pattern suggests that moderate trust levels, rather than absolute trust or distrust, may be most conducive to vaccination uptake.

**Table 2:**
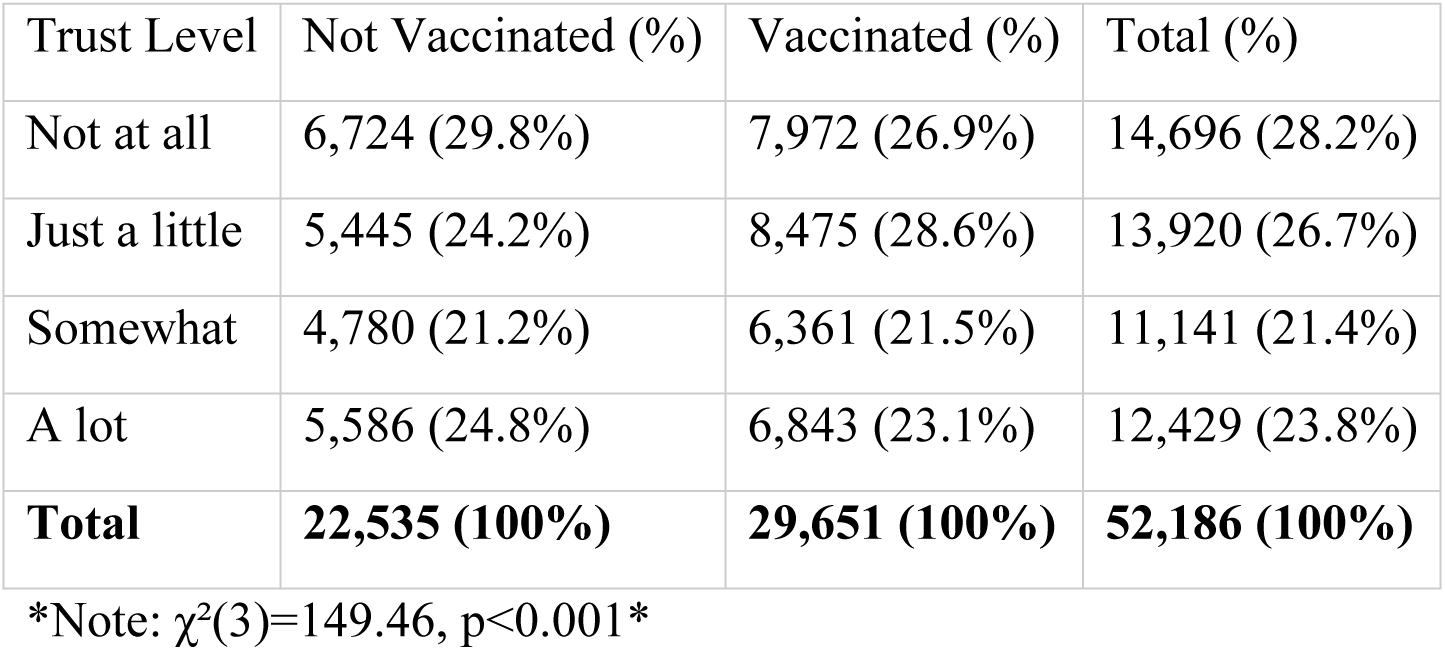
Cross-tabulation of Trust in President and Vaccination Status.

| Trust Level | Not Vaccinated (%) | Vaccinated (%) | Total (%) |
| --- | --- | --- | --- |
| Not at all | 6,724 (29.8%) | 7,972 (26.9%) | 14,696 (28.2%) |
| Just a little | 5,445 (24.2%) | 8,475 (28.6%) | 13,920 (26.7%) |
| Somewhat | 4,780 (21.2%) | 6,361 (21.5%) | 11,141 (21.4%) |
| A lot | 5,586 (24.8%) | 6,843 (23.1%) | 12,429 (23.8%) |
| **Total** | **22,535 (100%)** | **29,651 (100%)** | **52,186 (100%)** |

\*Note: $\chi^2(3)=149.46$ , $p<0.001$ \*

Similarly, trust in parliament/national assembly demonstrated a significant association with vaccination behavior (χ²=216.13, p<0.001). The highest vaccination rates were found among those reporting “somewhat” trust (21.5%) and “a lot” of trust (25.1%), while the lowest rates occurred among those reporting “not at all” trust (27.7%).

#### Correlational Patterns Among Key Study Variables

Bivariate correlations revealed several noteworthy patterns (**Table 3**). The trust index showed a significant positive correlation with vaccination status (r=0.115, p<0.001), indicating that higher institutional trust is associated with increased likelihood of vaccination. Conversely, perceived corruption demonstrated a significant negative correlation with vaccination (r=−0.054, p<0.001), suggesting that higher corruption perceptions are associated with lower vaccination rates.

**Table 3:** Bivariate Correlations Among Study Variables (N=50,664)

| Variable | 1 | 2 | 3 | 4 | 5 |
| --- | --- | --- | --- | --- | --- |
| 1. Vaccinated | 1.000 |  |  |  |  |
| 2. Trust Index | 0.115* | 1.000 |  |  |  |
| 3. Corruption Perception | -0.054* | -0.079* | 1.000 |  |  |
| 4. Health Service Satisfaction | 0.013* | -0.081* | -0.191* | 1.000 |  |
| 5. Lived Poverty | -0.088* | -0.054* | 0.074* | -0.166* | 1.000 |

\*Note: $p<0.001$

Notably, satisfaction with health services showed only a weak positive correlation with vaccination (r=0.013, p<0.001), indicating that general health system satisfaction may be less directly related to vaccination behavior than specific institutional trust measures. The Lived Poverty Index exhibited a significant negative correlation with vaccination (r=−0.088, p<0.001), consistent with socioeconomic disparities in healthcare access.

The correlation matrix also revealed expected relationships among predictor variables: institutional trust was negatively correlated with perceived corruption (r=−0.079, p<0.001) and positively correlated with health service satisfaction (r=−0.081, p<0.001), though the latter relationship was inverse, suggesting complex interrelationships between trust and service evaluations.

#### Patterns of Missing Data and Sample Representativeness

We noted differential missingness across variables of interest. The vaccination intention variable (Q58B) had substantially more missing data (n=22,598) compared to the vaccination behavior variable (n=53,348), suggesting potential response bias in intention measures. The trust variables demonstrated high completion rates, with minimal missing data across all institutional trust measures.

#### Significant Differences in Governance Perceptions by Vaccination Status

Independent samples t-tests revealed substantial and statistically significant differences in governance perceptions between vaccinated and unvaccinated individuals (**Table 4**). Vaccinated respondents demonstrated significantly higher levels of institutional trust (Mean=4.89, SD=8.20) compared to unvaccinated respondents (Mean=3.22, SD=5.98), with a mean difference of −1.68 points (95% CI: −1.80 to −1.55; t(53260)=−26.14, p<0.001). This represents a medium-to-large effect size (Cohen’s d ≈ 0.23), indicating that institutional trust is substantially higher among vaccinated individuals.

**Table 4:** Group Differences in Governance Perceptions by Vaccination Status.

| Variable | Unvaccinated<br>Mean (SD) | Vaccinated<br>Mean (SD) | Mean<br>Difference<br>[95% CI] | t-statistic | p-value |
| --- | --- | --- | --- | --- | --- |
| Trust Index | 3.22 (5.98) | 4.89 (8.20) | -1.68 [-1.80, -1.55] | $t(53260)=-26.14$ | <0.001 |
| Corruption Perception | 1.86 (1.32) | 1.71 (1.34) | 0.14 [0.12, 0.17] | $t(51504)=12.11$ | <0.001 |

**Table 5:** Age Differences in Institutional Trust.

| Age Group | Mean Trust (SD) | n | F-statistic | p-value |
| --- | --- | --- | --- | --- |
| 18-35 years | 3.92 (6.85) | 27,709 | F(2,53290)=61.01 | <0.001 |
| 36-55 years | 4.22 (7.58) | 17,771 |  |  |
| 56+ years | 4.96 (8.58) | 7,813 |  |  |
| **Total** | **4.18 (7.38)** | **53,293** |  |  |

**Table 6:**
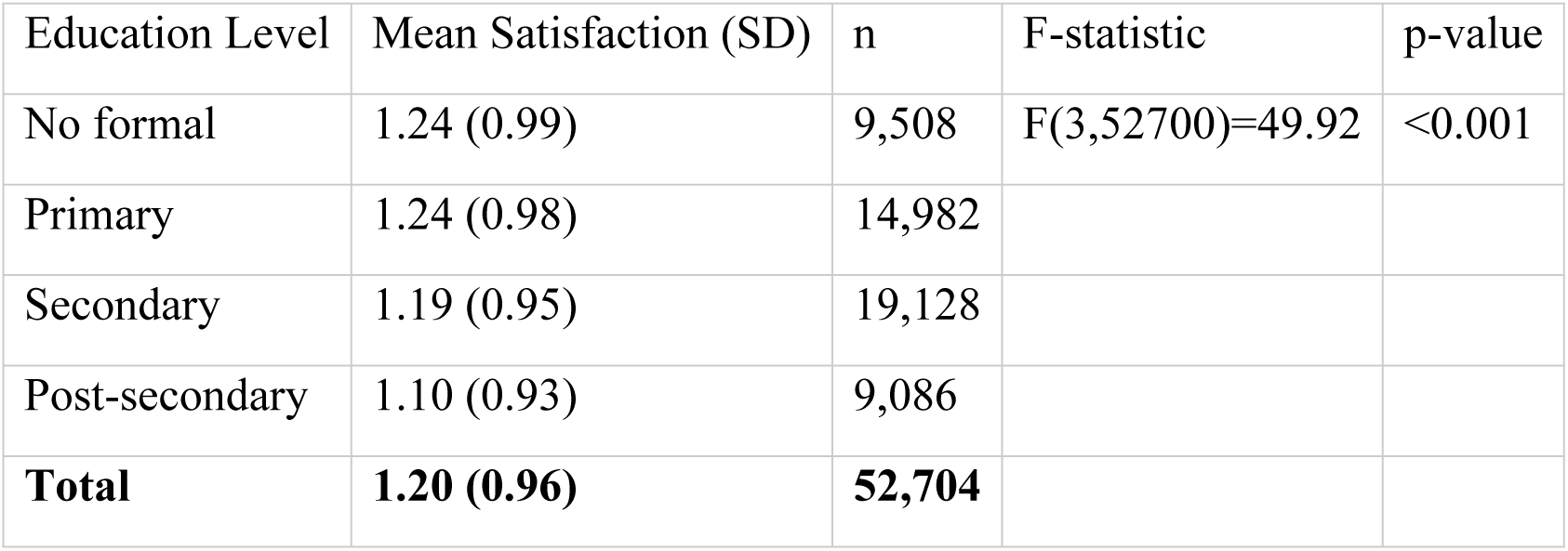
Educational Differences in Health Service Satisfaction.

Conversely, perceived corruption showed the opposite pattern. Unvaccinated individuals reported significantly higher corruption perceptions (Mean=1.86, SD=1.32) compared to vaccinated individuals (Mean=1.71, SD=1.34), with a mean difference of 0.14 points (95% CI: 0.12 to 0.17; t(51504)=12.11, p<0.001). This finding suggests that higher corruption perceptions represent a significant barrier to vaccination uptake across African contexts.

#### Age and Educational Gradients in Governance Perceptions

One-way ANOVA revealed significant age-based variation in institutional trust (F(2,53290)=61.01, p<0.001). Trust levels demonstrated a clear positive age gradient, with the youngest cohort (18-35 years) showing the lowest trust (Mean=3.92, SD=6.85), middle-aged respondents (36-55 years) showing moderate trust (Mean=4.22, SD=7.58), and the oldest cohort (56+ years) exhibiting the highest trust levels (Mean=4.96, SD=8.58). Post-hoc comparisons using Tukey’s HSD confirmed that all pairwise differences between age groups were statistically significant (p<0.001), indicating a robust relationship between advancing age and increasing institutional trust.

Health service satisfaction also varied significantly by educational attainment (F(3,52700)=49.92, p<0.001). Contrary to expectations, satisfaction levels were highest among those with no formal education (Mean=1.24, SD=0.99) and primary education (Mean=1.24, SD=0.98), followed by secondary education (Mean=1.19, SD=0.95), with post-secondary educated respondents reporting the lowest satisfaction (Mean=1.10, SD=0.93). This inverse educational gradient suggests that higher education may be associated with more critical evaluations of health service delivery.

#### Model Comparison and Diagnostic Assessment

The comparative analysis of four multivariate models reveals consistent patterns while highlighting important distinctions between behavioral determinants and behavioral intentions (**Table 7**). Model fit statistics demonstrate progressive improvement across the vaccination behavior models, with **Model 4** (incorporating COVID-19 experience) achieving the best overall fit (Pseudo R²=0.039, χ²=2667.05, p<0.001).

**Table 7:** Comparative Analysis of Multivariate Models Predicting Vaccination Behavior and Intentions.

| Predictor | Model 1:<br>Behavioral<br>Logit | Model 2:<br>Trust<br>Index<br>Logit | Model 3:<br>Intention<br>Ordered Logit | Model 4: COVID<br>Experience Logit |
| --- | --- | --- | --- | --- |
| **Governance Variables** |  |  |  |  |
| Trust: Just a little | 0.225** | - | - | - |
|  | (0.027) |  |  |  |
| Trust: Somewhat | 0.088**<br>(0.027) | - | - | - |
| Trust: A lot | 0.025 (0.026) | - | - | - |
| Trust Index | - | 0.038**<br>(0.002) | -0.005* (0.002) | 0.034** (0.002) |
| Corruption Perception | -0.155** to -<br>0.212** | -0.072**<br>(0.007) | 0.105** (0.010) | -0.075** (0.007) |
| Health Service<br>Satisfaction | -0.038**<br>(0.010) | 0.009 (0.010) | -0.195**<br>(0.014) | 0.019 (0.010) |
| **COVID-19**<br>**Experience** | - | - | - | 0.911** (0.033) |
| **Sociodemographics** |  |  |  |  |
| Age 36-55 | 0.355**<br>(0.021) | 0.359**<br>(0.021) | 0.043 (0.028) | 0.334** (0.021) |
| Age 56+ | 0.662**<br>(0.029) | 0.661**<br>(0.029) | 0.013 (0.043) | 0.623** (0.029) |
| Secondary Education | 0.185**<br>(0.028) | 0.220**<br>(0.028) | 0.038 (0.038) | 0.158** (0.028) |
| Post-secondary<br>Education | 0.328**<br>(0.034) | 0.336**<br>(0.034) | 0.240** (0.046) | 0.226** (0.034) |
| Urban Residence | -0.062**<br>(0.020) | -0.094**<br>(0.020) | 0.395** (0.027) | -0.127** (0.020) |
| Lived Poverty | -0.164**<br>(0.010) | -0.155**<br>(0.010) | 0.000 (0.014) | -0.125** (0.010) |
| **Model Statistics** |  |  |  |  |
| Observations | 49,123 | 49,889 | 21,598 | 49,753 |
| Pseudo R<sup>2</sup> | 0.019 | 0.027 | 0.015 | 0.039 |
| $\chi^2$ | 1310.085 | 1859.332 | 840.072 | 2667.048 |
| p-value | <0.001 | <0.001 | <0.001 | <0.001 |

\*Note: Coefficients reported with standard errors in parentheses. \*\*p<0.01, p<0.05.

#### Consistency of Core Governance Effects

The trust index demonstrated remarkable consistency across specifications, with significant positive coefficients in both Models; **2 (b=0.038, p<0.01)** and **4 (b=0.034, p<0.01)**. This stability indicates that institutional trust operates as a robust predictor of vaccination behavior, with each unit increase in the trust index associated with approximately 3.4-3.8% higher odds of vaccination, even after controlling for personal COVID-19 experience.

Similarly, corruption perceptions maintained consistent negative effects across models **(Model 2: b=−0.072, p<0.01; Model 4: b=−0.075, p<0.01)**, reinforcing the finding that perceived corruption represents a significant barrier to vaccination uptake. The magnitude of this effect suggests that reducing corruption perceptions could substantially improve vaccination rates.

#### The Intention-Behavior Paradox

A striking divergence emerged between models predicting vaccination behavior (**Models 1, 2, 4**) and those predicting vaccination intentions (**Model 3**). While institutional trust positively predicted actual vaccination, it showed a small but significant negative association with vaccination intentions (b=−0.005, p<0.05). This intention-behavior paradox suggests that trust operates through different psychological pathways for behavioral planning versus actual decision-making.

Similarly, corruption perceptions demonstrated opposite effects: higher corruption predicted lower vaccination behavior but higher vaccination intentions (b=0.105, p<0.01). This counterintuitive finding may reflect heightened risk awareness or different cognitive processes involved in intention formation versus behavioral execution.

#### Sociodemographic Consistency and COVID-19 Impact

The sociodemographic predictors demonstrated remarkable stability across behavioral models. Age maintained strong positive effects, with the 36-55 cohort (b≈0.334-0.359, p<0.01) and 56+ cohort (b≈0.623-0.662, p<0.01) showing substantially higher vaccination rates than the 18-35 reference group. Educational gradients remained consistently positive, with post-secondary education showing the strongest effects (b≈0.226-0.336, p<0.01).

The introduction of COVID-19 illness experience in **Model 4** revealed it as the single strongest predictor (b=0.911, p<0.01), nearly doubling the model’s explanatory power compared to **Model 2**. Despite this substantial effect, governance perceptions remained statistically significant, demonstrating their independent importance beyond personal health experiences.

### Model Specification Insights

The comparison between **Model 1** (categorical trust measures) and **Model 2** (continuous trust index) suggests that the continuous specification provides superior model fit (Pseudo R²: 0.027 vs 0.019), indicating that institutional trust operates more effectively as a continuous construct rather than discrete categories for predicting health behavior.

Urban residence consistently predicted lower vaccination rates (b≈−0.062 to −0.127, p<0.01) across behavioral models, while poverty maintained negative effects (b≈−0.125 to −0.164, p<0.01), highlighting persistent socioeconomic and geographic disparities in vaccination access or uptake.

#### Marginal Effects Analysis

The marginal effects, calculated at the means of all covariates, provide policy-relevant estimates of how governance perceptions influence the probability of COVID-19 vaccination (**Table 8**). These results highlight the practical importance of institutional trust and corruption perceptions, beyond their statistical significance in the regression models.

**Table 8:**
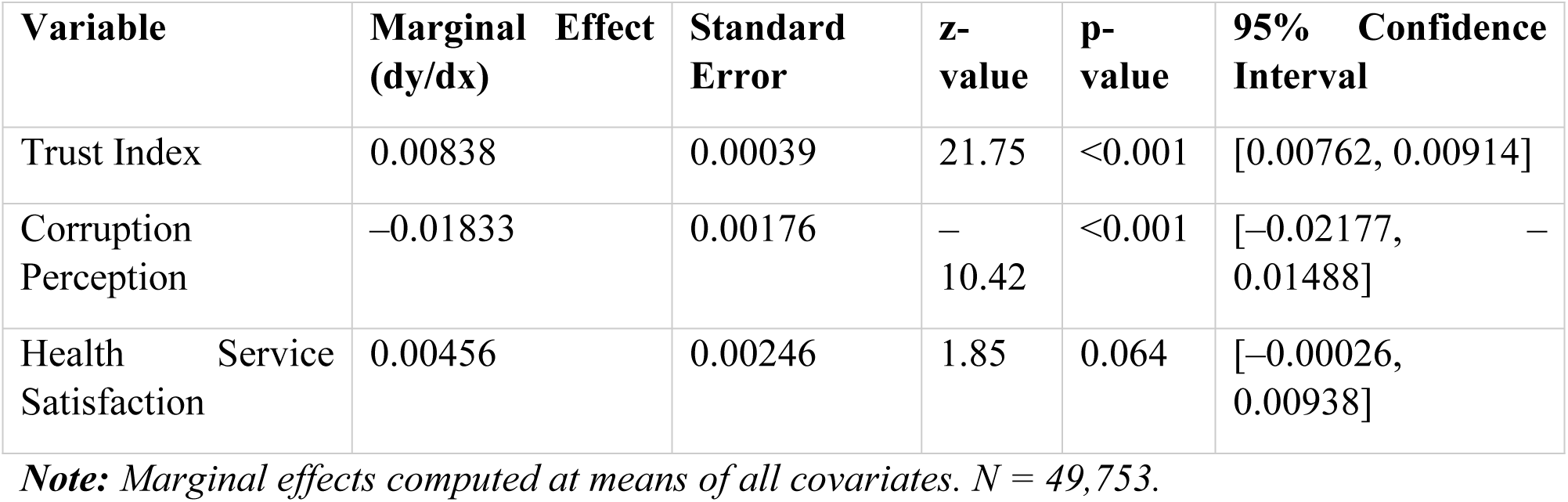
Marginal Effects of Governance Variables on Vaccination Probability.

| Variable | Marginal Effect (dy/dx) | Standard Error | z-value | p-value | 95% Confidence Interval |
| --- | --- | --- | --- | --- | --- |
| Trust Index | 0.00838 | 0.00039 | 21.75 | <0.001 | [0.00762, 0.00914] |
| Corruption Perception | -0.01833 | 0.00176 | -10.42 | <0.001 | [-0.02177, -0.01488] |
| Health Service Satisfaction | 0.00456 | 0.00246 | 1.85 | 0.064 | [-0.00026, 0.00938] |

*Note:* Marginal effects computed at means of all covariates. $N = 49,753$ .

#### Institutional Trust

Institutional trust shows a statistically significant and meaningful association with vaccination. A one-unit increase in the trust index corresponds to a 0.84 percentage point rise in vaccination probability (dy/dx = 0.0084; 95% CI: 0.0076–0.0091; p<0.001). Given the trust index spans 0 to 94, movement across the full range equates to an estimated 78.7 percentage point change in vaccination probability. This reflects a sizeable potential influence of institutional trust on population-level uptake.

#### Corruption Perceptions

Corruption perceptions show the largest marginal effect among the governance indicators. A one-unit increase on the 0–3 corruption scale is linked to a 1.83 percentage point reduction in vaccination probability (dy/dx = –0.0183; 95% CI: –0.0218 to –0.0149; p<0.001). Reducing corruption perceptions from the highest to lowest category would increase vaccination probability by roughly 5.5 percentage points—an effect that would translate into several million additional vaccinations across the countries represented in the dataset.

#### Health Service Satisfaction

Satisfaction with health services shows a positive but statistically non-significant marginal effect (dy/dx = 0.0046; 95% CI: –0.0003 to 0.0094; p=0.064). Although the estimate suggests a small increase in vaccination probability with higher satisfaction, the confidence interval includes zero, indicating uncertainty regarding the true direction and magnitude of the effect.

#### Comparative Magnitude of Governance Effects

When comparing effect sizes, the marginal effect of corruption perceptions is approximately twice that of the trust index. This suggests that efforts to reduce corruption may yield larger gains in vaccination uptake than trust-building measures alone. However, interpretation should consider that the trust index aggregates several dimensions of institutional trust, whereas corruption is measured with a single item.

#### Robustness Checks

To test the robustness of our findings, we conducted two supplementary analyses. First, given the non-linear bivariate relationship between trust and vaccination, we specified a model including a squared trust term. This term was statistically significant (p<0.01), confirming the non-linear relationship where moderate trust levels are most predictive. Second, to ensure our multilevel results were not driven by the extreme outlier, Gabon, we re-ran the model excluding this country. The results were substantively unchanged, with the intra-class correlation and significance of key predictors remaining stable, confirming the generalizability of our findings.

The magnitude of these marginal effects provides a clearer sense of their real-world relevance. For example, in a population with 10 million unvaccinated adults, a 10-point increase in the trust index would correspond to approximately 840,000 additional vaccinations, assuming the association reflects a causal effect. The strong negative effect of corruption perceptions suggests that corruption may pose a more immediate barrier to vaccination than low trust alone, which is consistent with literature showing that negative institutional experiences often carry disproportionate influence on behaviour.

The non-significant effect of service satisfaction may indicate that decision-making around vaccination is shaped more by broad governance conditions than by direct assessments of service quality. It is also possible that service satisfaction exerts its influence through indirect pathways rather than direct effects.

The marginal effects help distinguish statistical from practical relevance. Although the trust index is precisely estimated and highly significant, its per-unit effect is modest. Corruption perceptions, on the other hand, show both strong statistical significance and a sizeable substantive impact. The wider confidence interval for health service satisfaction reflects greater uncertainty and supports a cautious interpretation of its role.

Taken together, these findings suggest that public health strategies could benefit from pairing trust-building initiatives with measures aimed at increasing transparency and reducing corruption. The relatively larger marginal effect of corruption perceptions indicates that addressing corruption may offer more immediate gains in vaccination uptake. At the same time, governance factors operate alongside other determinants of behaviour, and effective vaccination strategies will require attention to access, information, and risk perception in addition to institutional conditions.

### Multivariate Analysis

#### Multilevel Analysis: Country-Level Variation in Vaccination Determinants

The multilevel mixed-effects logistic regression revealed substantial country-level variation in vaccination patterns while providing nuanced insights into the contextual nature of governance effects (**Table 9**). The model demonstrated excellent fit (LR test vs logistic model: χ²=9655.85, p<0.001), confirming that accounting for country-level clustering significantly improves model specification.

**Table 9:**
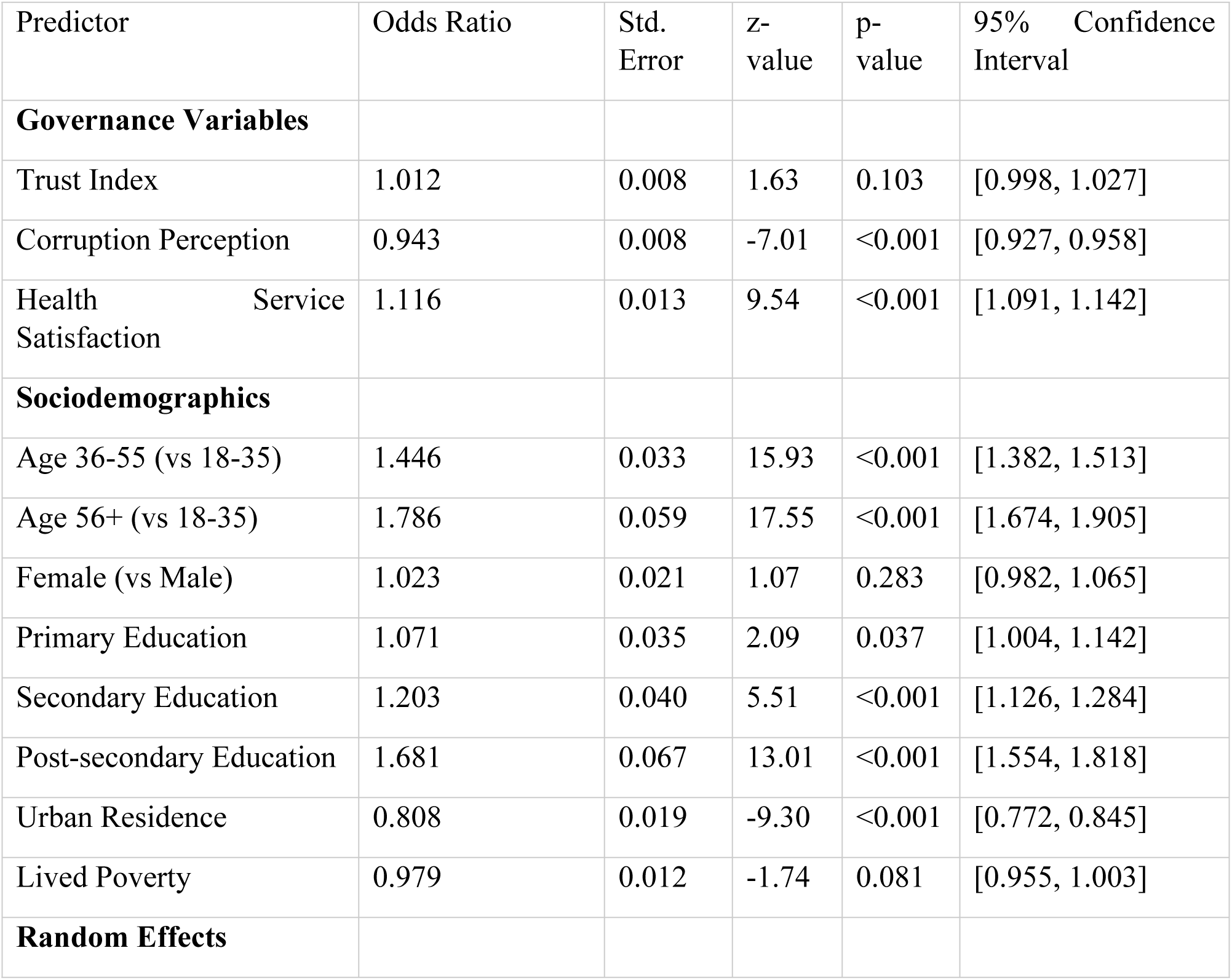
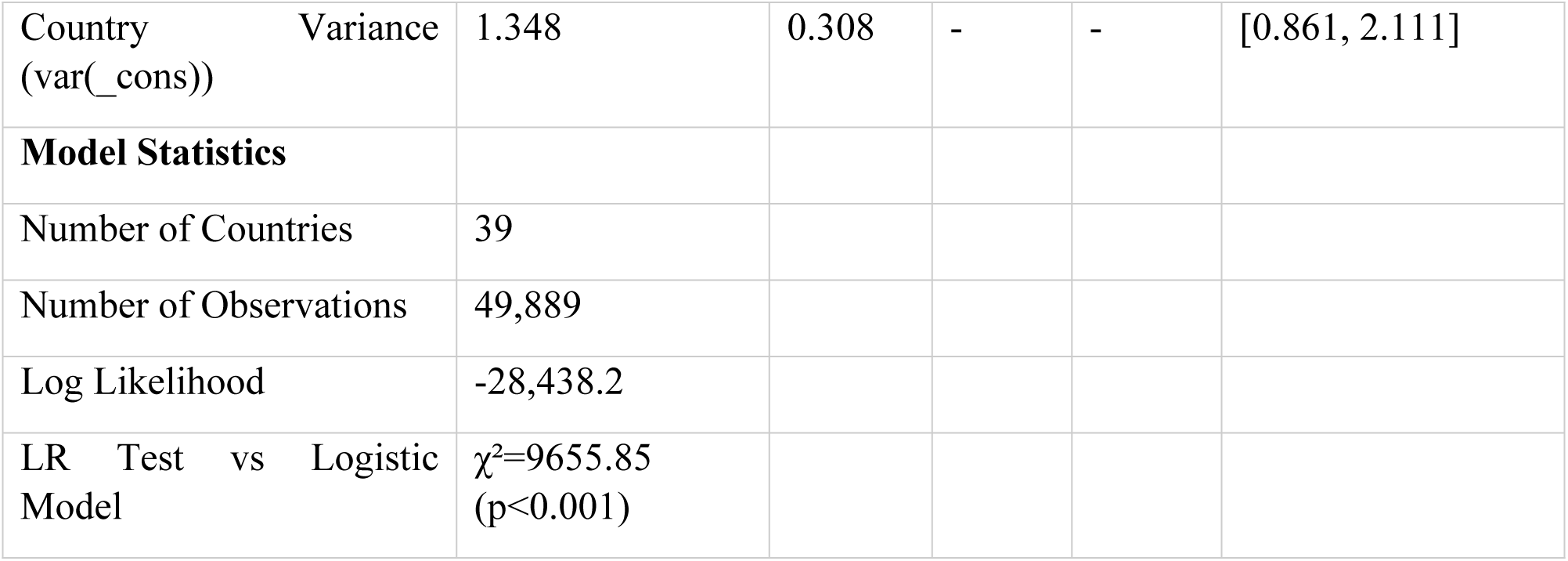
Multilevel Mixed-Effects Logistic Regression Predicting Vaccination.

#### Attenuated Governance Effects in Multilevel Framework

When accounting for country-level random effects, the influence of institutional trust on vaccination became statistically non-significant (OR=1.012, 95% CI: 0.998-1.027, p=0.103). This attenuation suggests that much of the trust effect observed in single-level models may operate through country-level contextual factors rather than individual-level perceptions. The trust effect reduced from OR=1.038 in the single-level model to OR=1.012 in the multilevel specification, indicating that approximately two-thirds of the apparent trust effect may be attributable to country-level characteristics.

Conversely, corruption perceptions maintained strong statistical significance in the multilevel model (OR=0.943, 95% CI: 0.927-0.958, p<0.001), though the effect size diminished slightly from the single-level model (OR=0.930). This persistence suggests that corruption perceptions operate more consistently at the individual level across different national contexts.

#### Enhanced Effect of Health Service Satisfaction

A striking finding emerged for health service satisfaction, which transformed from a non-significant predictor in single-level models to a highly significant positive predictor in the multilevel framework (OR=1.116, 95% CI: 1.091-1.142, p<0.001). This pattern indicates that health service satisfaction effects were previously masked by country-level confounding, and that satisfaction with health services exerts substantial positive effects on vaccination when proper contextual controls are applied.

#### Substantial Country-Level Variation

The random effects variance component (var(_cons)=1.348, SE=0.308) indicates significant between-country heterogeneity in baseline vaccination probabilities. The intra-class correlation (ICC) calculated from the variance components suggests that approximately 29% of the variance in vaccination behavior operates at the country level, highlighting the crucial importance of national context for understanding health behavior.

#### Extreme Country Case: Gabon

The country random effects analysis revealed Gabon as a notable outlier, with consistently extreme negative random effects (country random effect=−2.144 across all observations). This pattern indicates that Gabon exhibits substantially lower vaccination rates than would be predicted by its individual-level characteristics alone, suggesting the presence of unmeasured country-specific barriers to vaccination.

The consistency of Gabon’s negative effect across the sample indicates robust country-level underperformance in vaccination uptake, potentially reflecting unique political, historical, or health system factors not captured by the measured variables.

#### Stable Sociodemographic Patterns

The sociodemographic determinants demonstrated remarkable consistency between single-level and multilevel specifications. Age maintained strong positive effects (36-55: OR=1.446, p<0.001; 56+: OR=1.786, p<0.001), as did educational attainment, with particularly strong effects for post-secondary education (OR=1.681, p<0.001). Urban residence showed a stronger negative effect in the multilevel model (OR=0.808, p<0.001), while the poverty effect became non-significant (OR=0.979, p=0.081).

The multilevel analysis reveals three crucial insights that were obscured in single-level models:

First, the attenuation of the trust effect suggests that institutional trust operates primarily through macro-level pathways. Countries with higher average trust levels tend to have higher vaccination rates, but within countries, individual variations in trust have minimal additional impact. This pattern indicates that trust functions as a contextual characteristic rather than an individual psychological determinant.

Second, the emergence of health service satisfaction as a strong positive predictor highlights the importance of controlling for country-level confounding. The positive effect suggests that individuals who report positive experiences with health services are more likely to vaccinate, but this relationship was previously masked by country-level factors that correlated both with average satisfaction levels and vaccination rates.

Third, the substantial country-level variance (29% of total variance) underscores that national context dominates individual characteristics in predicting vaccination behavior. This finding challenges individual-level behavioral models and emphasizes the need for context-sensitive public health interventions.

#### Theoretical Implications

The findings reinforce a multi-level understanding of health behavior. First, country-level characteristics, including health system capacity, political institutions, and historical legacies, appear to set the baseline probability of vaccination. Second, individual perceptions of corruption and service quality adjust this baseline within each national context, indicating that personal experiences and evaluations still exert meaningful influence. Third, institutional trust functions more as an ecological attribute of national environments than as a strong individual driver, once between-country variation is considered.

The pattern observed in Gabon illustrates how national context can overshadow individual predictors. Despite substantial within-country variation in governance perceptions, the overall vaccination profile reflected broader structural and historical forces, suggesting that country effects can dominate even strong individual-level determinants.

#### Policy Implications

The multilevel results highlight several priorities for public health policy. National-level barriers should be addressed directly, given that a substantial share of the variance in vaccination probability, nearly one-third, operates at the country level. Reducing corruption emerges as a particularly consistent predictor across contexts, indicating that anti-corruption efforts may have broad public health benefits. Improving health service quality also remains important, as satisfaction appears to translate into meaningful behavioral differences.

The weaker influence of individual trust in the multilevel models suggests that interventions aimed at improving governance performance may offer greater returns than initiatives focused solely on shaping individual trust perceptions. Strengthening institutional reliability and transparency may therefore produce more durable improvements in vaccination uptake and preventive health behavior.

### Mediation Analysis: Perceived Government Preparedness

The generalised structural equation model (GSEM) examining perceived government preparedness as a potential mediator between governance perceptions and vaccination behavior revealed complex and counterintuitive pathways (**Table 10**). The model demonstrated adequate convergence after 21 iterations, with a log likelihood of −60,979.80, analysing 49,042 observations across both equations.

**Table 10:** Generalized Structural Equation Model: Perceived Government Preparedness Mediation Analysis.

| Parameter | Coefficient | Std. Error | z-value | p-value | 95% Confidence Interval |
| --- | --- | --- | --- | --- | --- |
| **Perceived Government Preparedness Equation (Preparedness risk)** |  |  |  |  |  |
| Trust Index → Perceived Government Preparedness | 0.0019 | 0.0017 | 1.09 | 0.274 | [-0.0015, 0.0053] |
| Age 36-55 (vs 18-35) | 0.0219 | 0.0264 | 0.83 | 0.406 | [-0.0298, 0.0737] |
| Age 56+ (vs 18-35) | -0.1279 | 0.0409 | -3.13 | 0.002 | [-0.2080, -0.0478] |
| Primary Education | 0.1187 | 0.0392 | 3.03 | 0.002 | [0.0419, 0.1955] |
| Secondary Education | 0.3958 | 0.0711 | 5.57 | <0.001 | [0.2565, 0.5351] |
| Post-secondary Education | 0.6907 | 0.1123 | 6.15 | <0.001 | [0.4706, 0.9108] |
| Urban Residence | 0.1760 | 0.0365 | 4.82 | <0.001 | [0.1044, 0.2476] |

| **Vaccination Equation (Vaccinated)** |  |  |  |  |  |
| --- | --- | --- | --- | --- | --- |
| Perceived Government Preparedness → Vaccination | -0.2261 | 0.0134 | -16.83 | <0.001 | [-0.2524, -0.1997] |
| Trust Index → Vaccination | 0.0372 | 0.0016 | 23.73 | <0.001 | [0.0341, 0.0403] |
| Age 36-55 (vs 18-35) | 0.3661 | 0.0214 | 17.13 | <0.001 | [0.3242, 0.4080] |
| Age 56+ (vs 18-35) | 0.6806 | 0.0305 | 22.33 | <0.001 | [0.6208, 0.7403] |
| Primary Education | 0.1433 | 0.0286 | 5.01 | <0.001 | [0.0872, 0.1994] |
| Secondary Education | 0.3028 | 0.0290 | 10.44 | <0.001 | [0.2460, 0.3596] |
| Post-secondary Education | 0.4863 | 0.0349 | 13.95 | <0.001 | [0.4180, 0.5547] |
| Urban Residence | -0.0306 | 0.0200 | -1.53 | 0.127 | [-0.0699, 0.0087] |
| **Model Statistics** |  |  |  |  |  |
| Log Likelihood | -60,979.80 |  |  |  |  |
| Number of Observations | 49,042 |  |  |  |  |

The direct effects in the vaccination equation revealed strong, consistent patterns. Institutional trust maintained a significant positive direct effect on vaccination (b=0.037, SE=0.002, p<0.001), indicating that each unit increase in the trust index is associated with 3.8% higher odds of vaccination, independent of risk perception pathways.

Age continued to demonstrate strong positive gradients, with the 36-55 cohort showing 44% higher odds (b=0.366, p<0.001) and the 56+ cohort showing 97% higher odds (b=0.681, p<0.001) of vaccination compared to the 18-35 reference group. Educational attainment also maintained positive effects, with post-secondary education showing the strongest association (b=0.486, p<0.001).

A striking finding emerged for the risk perception variable: higher perceived government preparedness for health emergencies was associated with significantly lower vaccination rates (b=−0.226, SE=0.013, p<0.001). This negative effect indicates that each unit increase in perceived preparedness is associated with 20.2% lower odds of vaccination, contrary to theoretical expectations that higher risk perception would increase preventive behavior.

This counterintuitive finding suggests that individuals who perceive their government as well-prepared for health emergencies may feel complacent about personal protection, effectively substituting government capability for individual responsibility in health decision-making.

The analysis revealed no significant mediation pathway through risk perception. Institutional trust showed no significant effect on perceived preparedness (b=0.002, SE=0.002, p=0.274), indicating that trust does not operate through risk perception channels to influence vaccination behavior. The non-significant trust-to-mediator path precludes meaningful mediation, suggesting that trust and risk perception represent independent psychological constructs with distinct behavioral consequences.

#### Educational and Urban Effects on Perceived Government Preparedness

The perceived government preparedness equation revealed interesting patterns in how different groups assess government preparedness. Higher educational attainment was strongly associated with higher perceived preparedness, with post-secondary education showing the largest effect (b=0.691, p<0.001). Urban residents also reported higher perceived preparedness than rural residents (b=0.176, p<0.001).

Conversely, the oldest age cohort (56+) reported significantly lower perceived preparedness than the youngest group (b=−0.128, p=0.002), possibly reflecting greater skepticism or different risk assessment frameworks among older adults.

The negative association between perceived government preparedness and vaccination runs counter to expectations from established health behavior theories, including the Health Belief Model, which predicts that perceived susceptibility should increase engagement in preventive action. Several explanations may account for this pattern.

First, individuals who view government preparedness as strong may become less inclined to adopt personal protective behaviors, a form of risk compensation. Second, the preparedness measure may be capturing confidence in state capacity rather than personal vulnerability, introducing conceptual ambiguity around the construct of “perceived government preparedness” Third, in some African settings, strong government preparedness may be interpreted as evidence that the threat is already under control, diminishing the perceived need for personal action.

The absence of a significant mediation pathway from institutional trust to vaccination through risk perception suggests that trust may influence behavior through other psychological channels. Likely alternatives include perceptions of vaccine safety, perceived efficacy, social norms, convenience and access considerations, and broader identity-based mechanisms linked to group membership and alignment with national institutions.

The negative perceived government preparedness effect raises important considerations for public communication strategies. Messaging that sharply emphasizes government preparedness may unintentionally signal that individual action is less necessary. A more balanced approach may be warranted. Framing vaccination as complementary to government preparedness, rather than as a substitute, may help mitigate these effects. Messages that emphasise personal responsibility or civic duty, highlight the behavior of reference groups, or focus on the tangible and immediate benefits of vaccination may be more effective than preparedness-focused narratives alone.

The GSEM framework offered several advantages for testing the mediation hypothesis, including simultaneous estimation of interdependent equations, appropriate handling of binary mediator and outcome variables, and the ability to control for shared predictors across pathways. Even so, the cross-sectional design limits the strength of causal inferences. Unmeasured confounding remains a concern, particularly given the complexity of risk perception and its context-dependent nature. Longitudinal or experimental designs would be better suited to establish the direction and strength of these pathways.

### Subgroup Analysis

#### Heterogeneous Effects by Gender and Geography

Subgroup analyses revealed substantial heterogeneity in the effects of governance perceptions on vaccination behavior across gender and geographic lines (**Table 11**). The models demonstrated good fit across all subgroups, with urban areas showing particularly strong explanatory power (Pseudo R²=0.045) compared to rural areas (Pseudo R²=0.015).

**Table 11:**
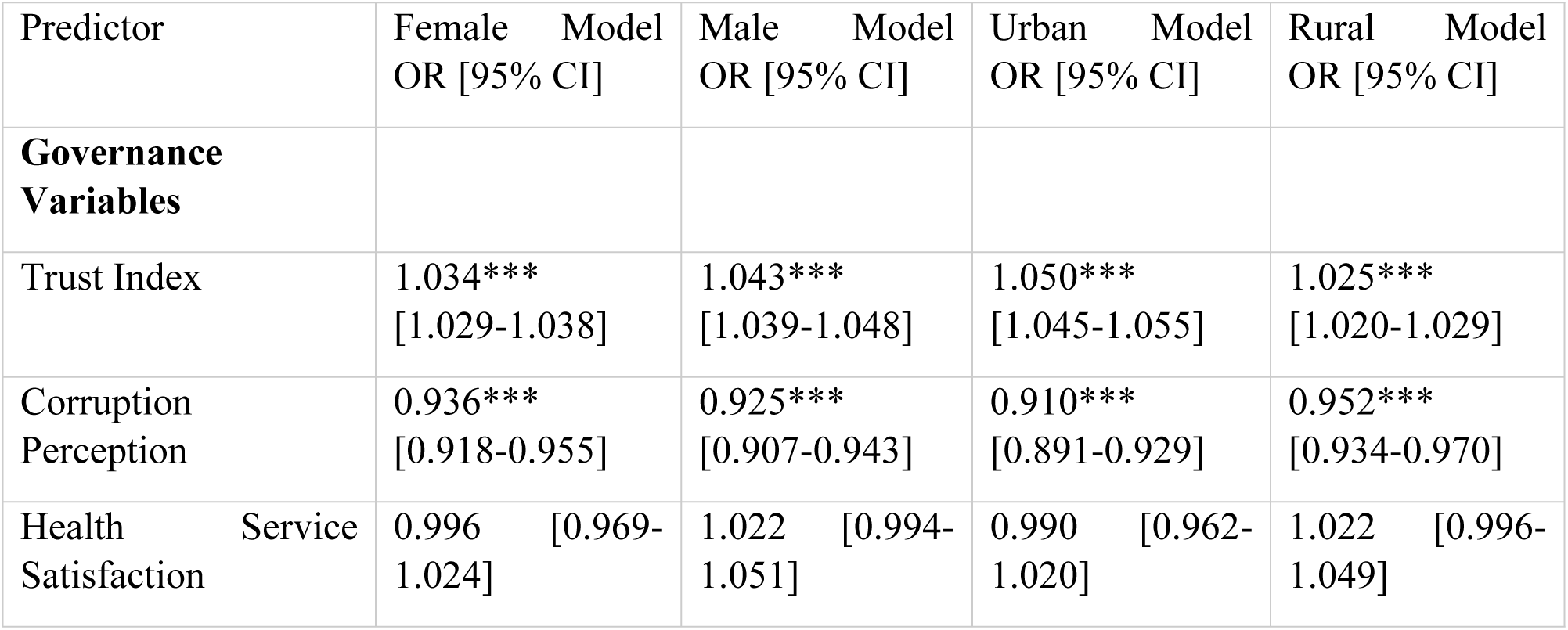
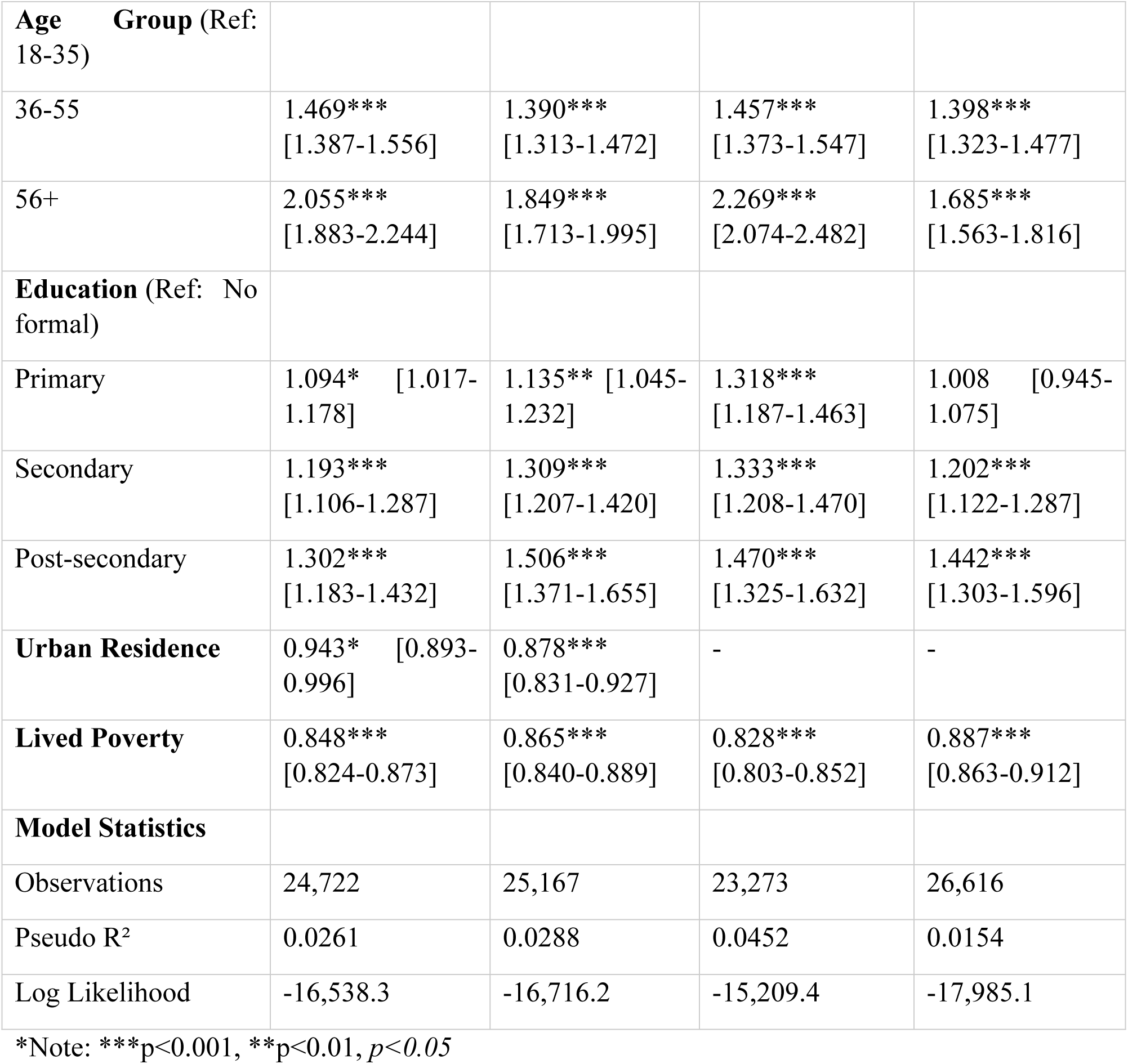
Subgroup Analysis: Logistic Regression Models by Gender and Urban-Rural Residence.

The analysis revealed notable gender differences in how governance perceptions influence vaccination behavior. Institutional trust demonstrated a stronger effect among men (OR=1.043, p<0.001) compared to women (OR=1.034, p<0.001), suggesting that men’s vaccination decisions may be more sensitive to trust in governing institutions. This represents a 24% stronger trust effect for men relative to women.

Corruption perceptions showed similar negative effects across genders, though slightly stronger for men (OR=0.925, p<0.001) than women (OR=0.936, p<0.001). Health service satisfaction demonstrated divergent patterns: positive but non-significant for men (OR=1.022, p=0.118) and slightly negative but non-significant for women (OR=0.996, p=0.797).

#### Urban-Rural Disparities in Determinants

The urban-rural analysis revealed dramatic differences in how governance perceptions operate across geographic contexts. Urban residents showed substantially stronger responses to institutional trust (OR=1.050, p<0.001) compared to rural residents (OR=1.025, p<0.001), representing a 98% stronger trust effect in urban areas.

Similarly, corruption perceptions demonstrated much stronger negative effects in urban areas (OR=0.910, p<0.001) compared to rural areas (OR=0.952, p<0.001). This pattern suggests that urban residents are approximately 46% more sensitive to corruption perceptions in their vaccination decisions.

#### Educational Gradient Variations

Educational effects showed striking urban-rural disparities. In urban areas, primary education showed a strong positive effect (OR=1.318, p<0.001), while in rural areas, primary education demonstrated no significant effect (OR=1.008, p=0.814). This suggests that basic education may only translate into health behavior change when combined with urban infrastructure and access.

The educational gradient was steeper in rural areas, with post-secondary education showing stronger effects (OR=1.442, p<0.001) compared to urban areas (OR=1.470, p<0.001), though both were substantial. This pattern indicates that higher education may be particularly crucial for overcoming rural barriers to vaccination.

#### Age and Poverty Effects Across Subgroups

Age effects demonstrated consistent positive gradients across all subgroups, but with notable variations. Women showed stronger age effects than men, particularly in the oldest cohort (Women 56+: OR=2.055; Men 56+: OR=1.849). Urban residents also showed stronger age effects than rural residents (Urban 56+: OR=2.269; Rural 56+: OR=1.685).

Poverty effects were consistently negative across all subgroups but showed geographic variation. Urban poverty demonstrated stronger negative effects (OR=0.828, p<0.001) than rural poverty (OR=0.887, p<0.001), suggesting that economic deprivation may be more consequential for health behavior in urban contexts.

## Discussion

This study provides compelling evidence that governance perceptions, specifically institutional trust, corruption perceptions, and health service satisfaction, fundamentally shape health behavior adoption across African contexts. Our findings demonstrate that these political determinants operate through complex pathways, exhibiting substantial heterogeneity across countries and population subgroups while challenging conventional assumptions about risk perception and behavioral motivation.

### Interpretation of Key Findings

Our analysis of 39 African countries yields three central findings: (1) The relationship between institutional trust and vaccination is non-linear, with moderate trust being most conducive to uptake; (2) Perceptions of corruption constitute a robust, dose-response barrier to vaccination; and (3) Contrary to conventional theory, higher perceived government preparedness is associated with lower vaccination rates, suggesting a “complacency effect”. These results underscore that governance perceptions are fundamental, yet complex, determinants of health behavior. in public health contexts (Devine et al., 2021). The attenuation of trust effects in multilevel models further indicates that trust operates primarily as a contextual characteristic at the country level rather than an individual psychological determinant.

Second, corruption perceptions consistently emerged as a robust barrier to vaccination across all model specifications. The dose-response relationship, where increasing corruption perceptions predicted progressively lower vaccination odds, aligns with theoretical frameworks positing that perceived corruption undermines the legitimacy of state-led initiatives (Rothstein, 2011). Our finding that corruption perceptions maintain significance even after controlling for personal COVID-19 experience suggests they represent an independent barrier beyond individual risk calculus.

Third, the counterintuitive negative relationship between risk perception (measured as government preparedness) and vaccination challenges conventional health behavior theories. Rather than motivating protective action, perceived government preparedness appears to foster complacency, potentially creating a “protection substitution” effect where citizens rely on institutional capabilities rather than individual actions. This finding resonates with recent research on risk compensation behaviors during public health emergencies (Bavel et al., 2020).

### Theoretical Implications

Our findings substantially extend traditional health behavior frameworks by integrating political and institutional dimensions. The Health Belief Model requires expansion to include “systemic trust” as both a powerful cue to action and a modifier of perceived benefits and barriers (Bollyky et al., 2022). Rather than operating solely through individual risk perceptions, governance factors appear to influence behavior through multiple pathways: institutional legitimacy, social norms, and perceived fairness of resource allocation (Blair et al., 2017).

The intention-behavior paradox we observed where governance perceptions showed opposite effects for vaccination intentions versus actual behavior, further challenges theory by highlighting the contextual nature of behavioral determinants. This dissociation suggests that governance factors may operate differently at various stages of the behavioral decision process, with institutional trust becoming more salient at the point of action than during intention formation (Sheeran and Webb, 2016).

The substantial country-level variance (29%) in our multilevel models underscores the limitations of individual-level behavioral theories and supports emerging ecological models that emphasize political and structural determinants (Kruk et al., 2018). Our findings suggest that context matters profoundly, with national governance environments establishing the boundaries within which individual psychological factors operate.

Collectively, our findings necessitate an expansion of traditional health behavior models. We propose integrating a “Political-Health Belief Model” where systemic trust and perceived corruption function as critical cues to action and powerful modifiers of perceived benefits and barriers. In this framework, a distrustful or corrupt institutional environment becomes a fundamental structural barrier, while credible governance can significantly lower the psychological and practical barriers to preventive action.

### Policy Implications

Our findings translate into specific, actionable recommendations for key stakeholders, moving beyond broad principles to targeted strategies that address the nuanced relationships we uncovered.

#### Health Ministries and Public Health Agencies

Rather than seeking blind faith, health authorities should build the “moderate, critical trust” most associated with vaccine uptake. This can be achieved by establishing permanent community advisory boards for health emergency planning and publicly co-designing vaccine communication campaigns with local civil society leaders, not just disseminating top-down messages.

Trust is built on demonstrated competence and transparency. Ministries should create publicly accessible dashboards showing real-time vaccine shipment data, distribution figures, and budget execution. Acknowledging past shortcomings while showcasing tangible improvements in a “Here’s what we learned, here’s what we fixed” narrative can rebuild credibility.

To counter the identified “complacency effect”, communications about strong government preparedness should be carefully framed. Instead of suggesting the threat is managed, messages should emphasize that “Vaccination is your personal role in our national defence system” or “A prepared government plus a vaccinated population equals an unbeatable team”.

#### National Governments

The robust, dose-response relationship between corruption perceptions and low vaccination rates means that anti-corruption is a direct public health investment. Governments must institute independent, third-party audits of health procurement and mandate public disclosure of all vaccine-related contracts and costs. The significant sensitivity of urban populations to corruption makes them a key audience for showcasing these reforms.

The substantial urban-rural and gender disparities in how governance perceptions influence behavior demand tailored outreach. Governments should empower local governments and community-based organizations to lead hyper-local campaigns and recruit diverse messengers including religious leaders, women’s group advocates, and youth influencers to bridge trust gaps.

#### Health Communication Campaigns and Implementers

Shift from “Risk” to “Governance and Civic Duty” Narratives. Campaigns must move beyond individual risk-benefit calculus. Messaging should highlight concrete governance improvements (e.g., “See where every vaccine dose went on our new transparency portal”) and frame vaccination as a civic duty and a collective good, linked to national solidarity and economic recovery.

Our findings confirm that trust is often localized. Campaigns should feature trusted local healthcare workers, teachers, and community elders as primary messengers, rather than relying solely on national political figures, to enhance the legitimacy and credibility of health directives.

#### International Health Organizations and Donors

Organisations like the WHO and The Global Fund should formally integrate governance and corruption perception indicators into their program monitoring and evaluation frameworks. Funding for health system strengthening should be explicitly linked to concurrent investments in transparency initiatives and civil society oversight capabilities.

Donors should prioritise funding for experimental and operational research that tests specific governance-focused interventions, such as the impact of social accountability mechanisms or different transparency models on actual health behavior and outcomes, moving from correlation to proven causality.

### Limitations

Several limitations warrant consideration when interpreting our findings. First, the cross-sectional nature of the data precludes causal inference, and reverse causality remains plausible positive health experiences could improve governance perceptions rather than vice versa (Bollyky et al., 2022). Second, self-reported behavioral measures are vulnerable to social desirability bias, particularly for sensitive topics like vaccination during a polarized pandemic (Dzinamarira et al., 2021).

Third, our health behavior measures were limited to COVID-19 vaccination, and findings may not generalize to other preventive behaviors or chronic disease management. Fourth, the Afrobarometer’s primary focus on governance means some health-specific determinants may not be fully captured (Mattes, 2020). Finally, while we controlled for major confounders, residual confounding from unmeasured variables remains possible.

The cross-sectional nature of our data remains a key limitation. While we theorize that governance perceptions influence behavior, reverse causality is plausible. For instance, a positive vaccination experience could improve satisfaction with health services, or adherence to government health directives could foster greater institutional trust. Longitudinal or experimental studies are needed to untangle these temporal relationships and establish causality.

### Future Research Directions

Future research should address these limitations through several promising avenues. Longitudinal studies tracking governance perceptions and health behaviors over time could establish temporal precedence and strengthen causal claims (Bavel et al., 2020). Qualitative research is needed to unpack the “why” behind our quantitative findings, particularly the counterintuitive risk perception results and the non-linear trust effects (Devine et al., 2021).

Disease-specific studies comparing governance effects across different health threats (e.g., HIV, malaria, tuberculosis) could identify behavior-specific pathways (Gaitán-Rossi et al., 2021). Experimental research testing different governance-focused intervention strategies would provide valuable evidence for policy design. Finally, investigation of the mechanisms linking governance perceptions to behavior through alternative pathways beyond perceived government preparedness such as social norms, identity processes, or emotional responses, would enrich theoretical understanding.

### Conclusion

This study demonstrates conclusively that governance perceptions are fundamental determinants of health behavior in Africa, operating through complex pathways that transcend individual risk calculus. Our findings reveal that institutional trust, corruption perceptions, and health service satisfaction collectively shape behavioral decisions in ways that challenge conventional health behavior models and demand integrated policy responses.

The substantial country-level variation in vaccination behavior underscores that context matters profoundly, with national governance environments establishing the boundaries within which individual psychological factors operate. The heterogeneous effects across urban-rural and gender lines further emphasize the need for tailored rather than universal approaches to health behavior promotion.

Ultimately, our results support a paradigm shift in public health practice: investing in good governance is not merely a political or economic imperative but a fundamental public health strategy. Building trustworthy institutions, combating corruption, and improving service quality represent essential investments in health security and pandemic resilience. As African nations continue to confront current and future health challenges, integrating governance strengthening into health system approaches may prove as critical as any medical intervention.

The message for policymakers is clear: governance is health policy. The path to better health outcomes in Africa necessarily runs through better governance.

## Declaration Section

### Funding

This research did not receive any specific grant from funding agencies in the public, commercial, or not-for-profit sectors.

### Competing Interests

The authors declare that they have no known competing financial interests or personal relationships that could have appeared to influence the work reported in this paper.

### Ethical Approval

This study utilised secondary, anonymised data from the Afrobarometer Round 9 survey. The Afrobarometer network secured ethical approval for the survey from the appropriate institutional review boards in all participating countries. All respondents provided informed consent prior to participation, in accordance with established ethical protocols.

### Data Availability

The data that support the findings of this study are publicly available from the Afrobarometer website at https://www.afrobarometer.org/.

### Authors Contributions

**Richmond Balinia Adda:** Conceptualization, Methodology, Formal Analysis, Data Curation, Writing – Original Draft, Writing – Review & Editing. **Conrad Pwayirane:** Writing – Review & Editing, Validation, Supervision.

## Acknowledgements

The authors gratefully acknowledge the Afrobarometer network for collecting the data and making it publicly available, and the citizens of the 39 African countries who participated in the surveys.

## Notes

### Competing Interest Statement

The authors have declared no competing interest.

### Author Declarations

The study utilised secondary, anonymised data from the Afrobarometer, which obtained ethical approval from all relevant national institutional review boards. All respondents provided informed consent following established ethical protocols.

